# Hidden risk in normal myocardial perfusion scans: AI-detected proximal coronary calcium on CT attenuation maps improves prognosis

**DOI:** 10.64898/2026.04.14.26350808

**Authors:** Jianhang Zhou, Robert JH Miller, Aakash Shanbhag, Aditya Killekar, Donghee Han, Krishna K Patel, Konrad Pieszko, Jirong Yi, Meghana Kiran Urs, Giselle Ramirez, Mark Lemley, Paul B. Kavanagh, Joanna X Liang, Assiata Kamagate, Valerie Builoff, Andrew J Einstein, Attila Feher, Edward J Miller, Albert J Sinusas, Terrence D Ruddy, Stacey Knight, Viet T Le, Steve Mason, Panithaya Chareonthaitawee, Samuel Wopperer, Erick Alexanderson, Isabel Carvajal-Juarez, Thomas L Rosamond, Leandro Slipczuk, Mark I. Travin, René R. S. Packard, Wanda Acampa, Mouaz Al-Mallah, Robert A deKemp, Ronny R. Buechel, Daniel S Berman, Damini Dey, Marcelo F Di Carli, Piotr J Slomka

## Abstract

**Purpose:** Spatial distribution of coronary artery calcium (CAC) may provide additional prognostic value in patients undergoing SPECT and PET myocardial perfusion imaging (MPI). We aimed to automatically identify CAC in proximal segments from attenuation correction CT (CTAC) scans using artificial intelligence (AI) and to evaluate prognostic significance in two large international multicenter registries.

**Methods:** From hybrid MPI/CT imaging (N=43,099) across 15 sites, we included 4,552 most relevant patients with 1) no prior coronary artery disease; 2) AI-derived mild CAC scores (1-99); and 3) normal perfusion (stress total perfusion deficit <5%). The independent associations between AI-identified proximal CAC and major adverse cardiovascular events (MACE) and all-cause mortality (ACM) were evaluated using multivariable Cox regression, likelihood ratio test (LRT), and continuous net reclassification index (NRI).

**Results:** Among the patients with mild CAC and normal perfusion (mean age 65±12 years, 51% male), 1,730 (38%) had proximal CAC. Over 3.6 (inter-quartile interval 2.1, 5.2) years follow-up, 599 (13%) and 444 (10%) patients had MACE or ACM, respectively. Proximal CAC was associated with an increased risk of MACE (adjusted hazard ratio [HR] 1.24, 95% CI 1.03–1.48, P=0.02) and ACM (adjusted HR 1.25, 95% CI 1.01–1.53, P=0.04) after the adjustment of CAC score and density, clinical risk factors, and perfusion deficit. Proximal CAC improved the risk stratification of MACE (LRT P=0.02; NRI 12%) and ACM (LRT P=0.04; NRI 12%).

**Conclusion:** In patients with mild CAC and normal perfusion, AI detection of proximal CAC identified a higher-risk group for adverse outcomes, highlighting its prognostic utility.

**Graphical Abstract:** 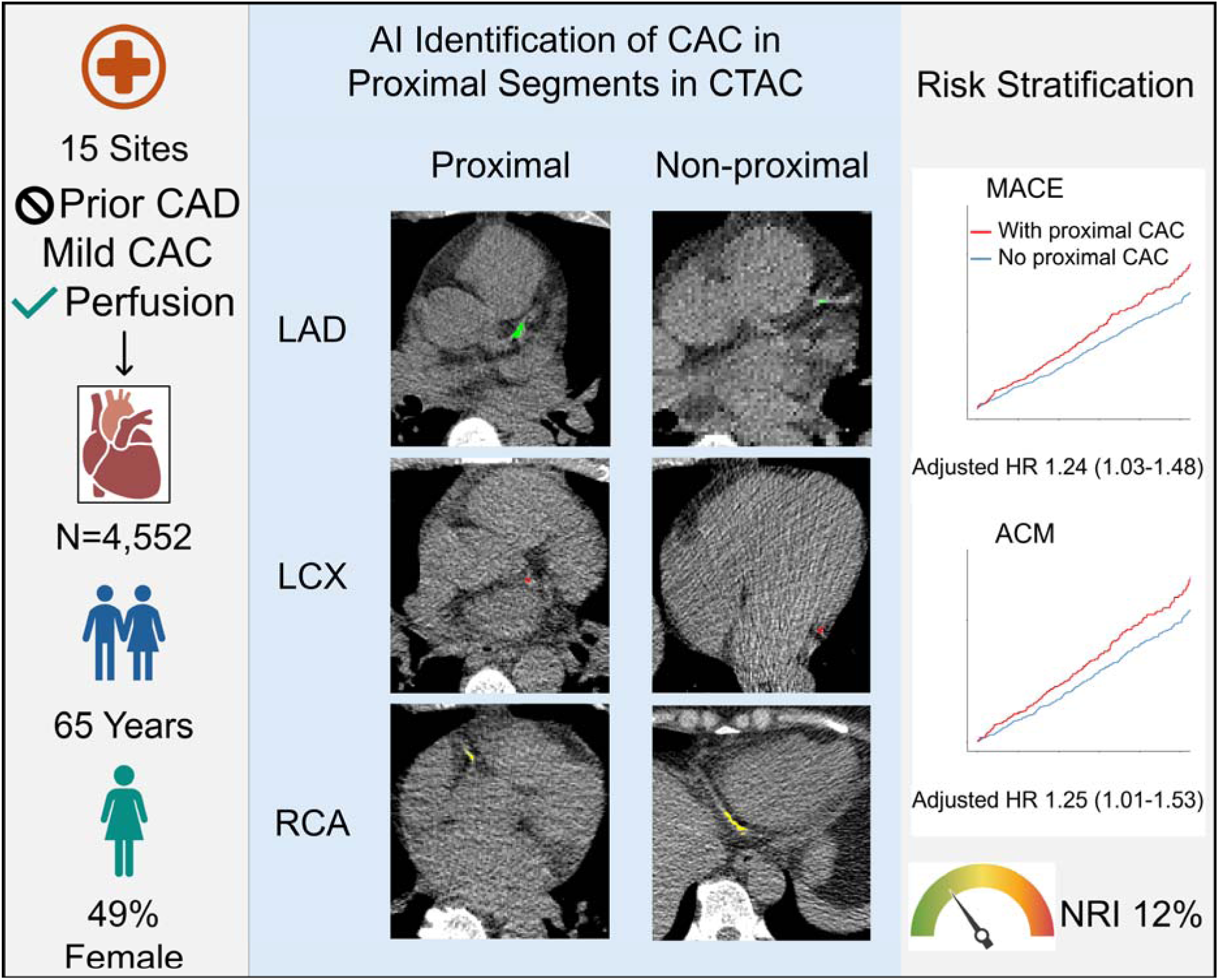

From patients who underwent hybrid myocardial perfusion imaging (MPI) from 15 sites, we analyzed those without prior coronary artery disease (CAD), mild coronary artery calcium (CAC) scores (1-99), and normal perfusion (stress total perfusion deficit <5%). A previously developed AI model was used to identify CAC lesions in proximal coronary segments on CT attenuation correction maps (CTAC). We evaluated associations with major adverse cardiovascular events (MACE) and all-cause mortality (ACM), showing risk stratification of proximal CAC and improvement by net reclassification index (NRI). CAC lesion color: green, left anterior descending artery (LAD) with left main artery; red, left circumflex artery (LCX); yellow, right coronary artery (RCA). Adjusted hazard ratios (HRs) are shown with 95% confidence intervals.

## Introduction

Coronary artery calcium (CAC) evaluation is a critical anatomic tool for the assessment of CAD that can benefit patients undergoing hybrid myocardial perfusion imaging (**MPI**)[1], including single-photon emission computed tomography (**SPECT**)[2, 3] and positron emission tomography (**PET**)[4, 5]. CAC assessment can improve the detection of obstructive coronary artery disease (**CAD**)[6–9]. Several large studies have demonstrated that the CAC score is an independent and complementary predictor of cardiovascular risk in patients undergoing MPI[10–17].

Beyond the CAC score as a single measure, CAC location in the arteries may further improve the diagnostic and prognostic value of CAC phenotyping for MPI patients[18]. In particular, proximal plaques are more prone to rupture, leading to coronary thrombosis[19, 20]. As a result, the presence of proximal coronary plaque is associated with a higher cardiovascular risk in patients undergoing coronary computed tomography (**CT**) angiography[21, 22] or CAC CT scans[23]. In asymptomatic adults with mild CAC, proximal CAC showed independent prognostic value in predicting MACE[23], However, it is unknown whether the CAC location derived from low-dose, ungated CT attenuation correction (**CTAC**) scans (routinely acquired with hybrid imaging) provides independent prognostic value in a similar patient group [10, 12, 24]. Artificial intelligence (**AI**) has been applied to CTAC for CAC scoring [16, 25–28] and integrated risk prediction[14, 15, 29]. However, on these ultra-low-dose images, it can be difficult to accurately identify all CAC plaques or determine their exact location.

Previously, we developed a novel AI algorithm that integrates CAC and cardiac tissue segmentation to automatically classify CAC lesions into proximal and non-proximal categories[30]. In this study, we evaluated the prognostic significance of CAC proximal involvement for MPI patients with mild CAC and normal perfusion – the group who may benefit most from further risk stratification by proximal CAC- in two large international multicenter registries comprising 15 sites.

## Material and Methods

### Study population

Consecutive patients with CTAC scans from hybrid SPECT/CT imaging (4 sites, N=10,989) and PET/CT imaging (13 sites, N=32,110; together 15 sites, 2 sites in both registries, N=43,099) of two international multi-center registries[31–33] were studied. The registry protocols were approved by the Institutional Review Board at all participating sites, and sites either obtained written informed consent or a waiver of consent for the use of the deidentified data. The overall registries were approved by the investigational review board at Cedars-Sinai Medical Center and complied with the Declaration of Helsinki. Clinical variables collected at each site were de-identified and anonymized prior to transfer to the core site using dedicated software compliant with the Health Insurance Portability and Accountability Act (HIPAA). Patients with a history of coronary artery disease (defined as previous myocardial infarction, percutaneous coronary intervention, or coronary artery bypass grafts; N=13,364) were excluded. Segmentation failure occurred in 63 patients (0.2% of 29,735) who were excluded. After exclusions, we had 29,672 patients.

### CT acquisition

All CT scans were acquired without contrast enhancement or cardiac gating. Details of acquisition protocols are shown in **Supplement Table 1** for SPECT/CT and **Supplement Table 2** for PET/CT.

### Perfusion imaging quantification

Stress total perfusion deficit (**TPD**) was used to determine perfusion status. Patients with a stress TPD < 5% were considered normal, and those with a value ≥ 5% were considered abnormal[34, 35]. Stress TPD was quantified using Quantitative Perfusion SPECT (QPS)/Quantitative Gated SPECT (QGS) software (Cedars-Sinai Medical Center, Los Angeles, California) [35, 36] for SPECT and the standard clinical quantitative PET package QPET (Cedars-Sinai Medical Center, Los Angeles, California) [37, 38] for PET.

### Automated coronary artery calcium segmentation

A previously validated deep-learning model was used to segment vessel-specific CAC lesions (left anterior descending [**LAD**], left circumflex [**LCX**], and right coronary arteries [**RCA**]) [27, 39–41]. Lesions in the left main artery (LM) were included in LAD. Total CAC scores were determined from the CAC segmentation on CTAC scans using the Agatston method [42] and the standard threshold of 130 Hounsfield Units [27, 39, 43].The scores in the Agatston Unit (AU) of 1-99 were categorized as mild. CAC density was computed and adjusted to slice thickness[44].

### Identification of proximal CAC lesions

A previously reported AI algorithm [30] was used to identify CAC lesions in the proximal coronary segments. The algorithm used cardiac tissue segmentations [45] and analyzed distances between each CAC lesion from the previous step and a reference point automatically placed on the ascending aorta. On the slice of the CT scan showing the base of the left ventricle, the pixel in the ascending aorta with the shortest distance to the left ventricle was set as the reference point. The reference point was set as a proxy for the left ostium. In short, proximal CAC detection was performed by XGBoost (eXtreme Gradient Boosting) [46] integrating: (i) distance from the lesion to the reference point; (ii) location of the lesion defined as the center of mass in absolute image coordinates (coronal, sagittal, axial axes); (iii) location of the reference point in absolute image coordinates; (iv) the coronary artery having the lesion (LAD, LCX, or RCA). The algorithm was trained on dedicated CAC CT scans of 303 asymptomatic adults. Proximal location of CAC in each coronary artery was determined based on the branches (LAD based on the first diagonal branch, LCX based on the first obtuse marginal branches, RCA based on the distance from the ostium to the origin of the first acute marginal artery [23, 47]. Proximal segments were defined as proximal LAD (including LM), proximal LCX, and proximal RCA. The algorithm was adapted to CTAC by replacing image coordinates with physical distances from the reference point to the lesion along the three imaging axes.

### Prognostic evaluation

The primary endpoint was major adverse cardiovascular events (**MACE**), defined as all-cause mortality, revascularization (percutaneous coronary intervention or coronary artery bypass grafting), admission for unstable angina, or myocardial infarction. The secondary endpoint was all-cause mortality. Cumulative event rates were computed with the Kaplan-Meier method. Survival curves were compared with the log-rank test. The multivariable Cox analysis included site, log(CAC+1), CAC density, and clinical factors (age, sex, body mass index [**BMI**], hypertension, dyslipidemia, diabetes, family history of heart disease, and current smoker) as covariates, along with stress TPD. Site was included in the adjustment to account for underlying shared frailty in patient demographics, clinical profiles, and imaging parameters. The proportional hazard assumption was assessed with the Schoenfeld residual test. No violation was found. Hazard ratios (**HRs**) were presented with 95% confidence intervals (**CIs**).

To evaluate the incremental discriminative value of proximal CAC in such patient groups, the proximal involvement was added to a base Cox multivariable model as a nested model. The base model included scan sites, CAC score and density, clinical risk factors, and stress TPD. The prediction performance of the two models was assessed by Harrell’s C-index and compared using the likelihood ratio test. The incremental value was estimated by the continuous net reclassification index (**NRI**)[48].

### Statistics

Statistical analyses were performed using R (Version 4.4.1, R Foundation for Statistical Computing, Vienna, Austria) and Python 3.7.16. Continuous variables are presented as mean ± standard deviation for normally distributed data and median (interquartile interval [IQI]) for non-normally distributed data. A two-tailed p-value < 0.05 was considered statistically significant for the statistical tests mentioned above.

## Results

### Patient characteristics

In total, 18,986 (64% of 29,672) patients had normal perfusion (stress TPD <5%). The deep learning model identified 17,830 (60% of 29,672) patients with CAC, of whom 6,691 had mild CAC (1-99). Together, we recognized 4,552 patients with mild CAC and normal perfusion for subsequent analyses (**Supplement Figure 1**). Among these patients, the algorithm for proximal lesion detection determined 1,730 (38%) patients having proximal CAC. The baseline characteristics for patients with and without proximal CAC are summarized in **Table 1**. Patients with proximal CAC were significantly older (mean age 67 years versus 63 years), had a lower BMI (mean 29 kg/m^2^ versus 31 kg/m^2^), and were more likely to have dyslipidemia (64% versus 60%), but less likely to have diabetes (27% versus 30%). Patients with proximal CAC had higher total CAC scores (median 39 AU versus 22 AU), lower non-proximal CAC scores (median 0 versus 22 AU), and denser calcification (2.0 versus 1.8). No significant difference was found in sex, hypertension, family history of CAD, or stress TPD.

**Table 1.** Baseline demographic and clinical characteristics by CAC proximal involvement.

|  | **With proximal**<br>**CAC** | **No proximal**<br>**CAC** | **P-value** |
| --- | --- | --- | --- |
| Number of patients | 1,730 | 2,822 |  |
| Age, years | 67 ± 11 | 63 ± 12 | < 0.001 |
| Male | 886 (51%) | 1,452 (51%) | 0.9 |
| Body mass index, kg/m<sup>2</sup> | 29 ± 6 | 31 ± 7 | < 0.001 |
| Hypertension | 1,212 (70%) | 1,939 (69%) | 0.3 |
| Diabetes | 459 (27%) | 851 (30%) | 0.009 |
| Dyslipidemia | 1,106 (64%) | 1,687 (60%) | 0.005 |
| Family history of CAD | 461 (27%) | 708 (25%) | 0.2 |
| Current Smoker | 413 (24%) | 600 (21%) | 0.040 |
| Stress TPD, % | 1.6 (0.6, 2.9) | 1.7 (0.7, 2.9) | 0.4 |
| CAC score, AU | 39 (19, 65) | 22 (11, 47) | < 0.001 |
| Proximal CAC score, AU | 23 (12, 43) | 0 | < 0.001 |
| Non-proximal CAC score, AU | 0 (0, 19) | 22 (11, 47) | < 0.001 |
| CAC density | 2.0 (1.6, 2.4) | 1.8 (1, 2) | <0.001 |
| Follow-up duration, years | 3.5 (2.0, 5.1) | 3.8 (2.1, 5.3) | 0.001 |
| MACE | 251 (15%) | 348 (12%) | 0.035 |
| All-cause mortality | 185 (11%) | 259 (9%) | 0.094 |

Continuous variables are presented as mean ± standard deviation or median (lower quartile, upper quartile). Categorical variables are presented as counts (percentages).
AU, Agatston unit; CAC, coronary artery calcium; CAD, coronary artery disease; MACE, major adverse cardiovascular events; TPD, total perfusion deficit.

### Prognostic values of proximal involvement

During the follow-up period of a median of 3.6 (IQI 2.1, 5.2) years, 599 (13%) patients had MACE, and 444 (10%) died. The risk stratification by proximal involvement is shown in **Figure 1**. Patients with proximal CAC had significantly higher cumulative MACE (P = 0.001) and all-cause mortality (P = 0.005).

**Figure 1.**
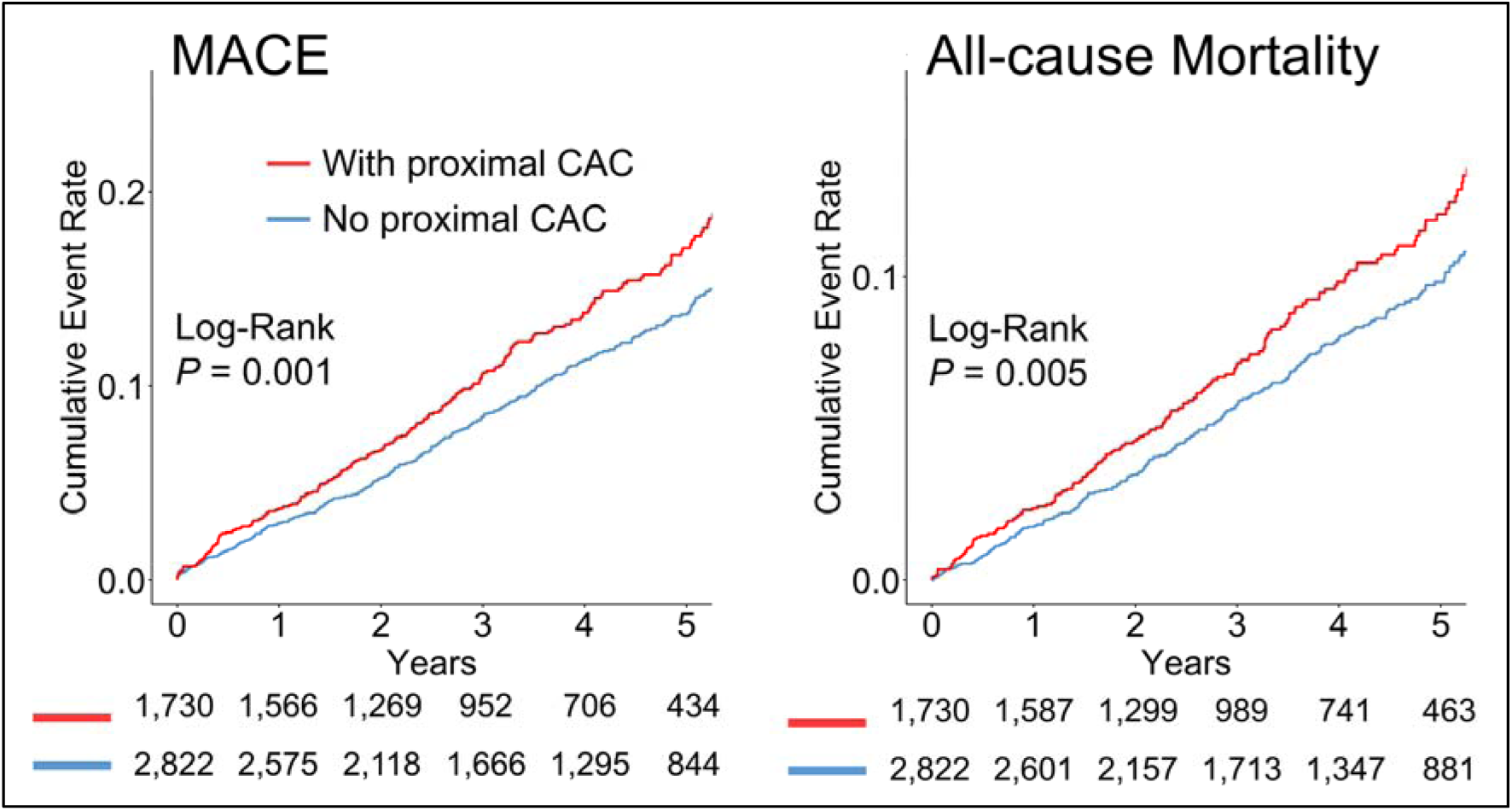
Kaplan-Meier survival curves stratified by the proximal involvement. Kaplan-Meier curves showing cumulative rates of major adverse cardiovascular events (MACE; left) and all-cause mortality (right). P-values from the log-rank test were shown.

The multivariable analyses are summarized in **Table 2**. After accounting for the total CAC score and imaging site, proximal CAC was associated with an increased risk of MACE (adjusted HR 1.38, 95% CI 1.15 – 1.64, P < 0.001) and all-cause mortality (adjusted HR 1.44, 95% CI 1.18 – 1.77, P < 0.001). Additional adjustment of CAC density gave the same adjusted HRs for both outcomes. After further adjustment of age, sex, BMI, and medical history, the increased risk of proximal CAC remained significant for MACE (adjusted HR 1.23, 95% CI 1.03 – 1.48, P = 0.020) and all-cause mortality (adjusted HR 1.24, 95% CI 1.01 – 1.53, P = 0.040). The significance was retained after accounting for stress TPD from perfusion imaging (MACE: adjusted HR 1.24, 95% CI 1.03 – 1.48, P = 0.020; all-cause mortality: adjusted HR 1.25, 95% CI 1.01 – 1.53, P = 0.036).

**Table 2.** Multivariable Cox regression of outcomes and CAC proximal involvement.

| Outcome | Model | Hazard ratio | 95% CI | P value |
| --- | --- | --- | --- | --- |
| MACE | Model 1 | 1.38 | 1.15-1.64 | <0.001 |
|  | Model 2 | 1.37 | 1.15-1.64 | <0.001 |
|  | Model 3 | 1.23 | 1.03-1.47 | 0.023 |
|  | Model 4 | 1.24 | 1.03-1.48 | 0.020 |
| ACM | Model 1 | 1.44 | 1.18-1.77 | <0.001 |
|  | Model 2 | 1.45 | 1.18-1.78 | <0.001 |
|  | Model 3 | 1.24 | 1.01-1.53 | 0.040 |
|  | Model 4 | 1.25 | 1.01-1.53 | 0.036 |

Model 1: proximal involvement, total CAC score, site
Model 2: Model 1 plus CAC density
Model 3: Model 2 plus age, sex, body mass index, hypertension, diabetes, dyslipidemia, family history of coronary heart disease
Model 4: Model 3 plus stress total perfusion deficit
Adjusted hazard ratios with 95% confidence intervals (CIs) are from Cox multivariable models. ACM, all-cause mortality; CAC, coronary artery calcium; MACE, major adverse cardiovascular events.

The incremental prognostic value of proximal involvement was outlined in **Supplement Table 3**. The base model reached a C-index of 0.645 ± 0.013 for the association with MACE. The nested model, including the proximal involvement, increased the C-index to 0.648 ± 0.013 (P = 0.020). In the continuous NRI analysis over a 4-year interval, proximal CAC measurements resulted in an overall improvement of 12% (95% CI 2%-19%) of MACE cases. For all-cause mortality, proximal involvement improved the C-index from 0.680 ± 0.015 to 0.683 ± 0.014 (P = 0.037) with an overall NRI of 12% (95% CI 1%-24%).

Case illustrations are provided in **Figure 2**. The top panel shows two male patients in their 50s with similar CAC scores and densities, and stress TPD both as 0. The patient with CAC in the proximal RCA had MACE 1.7 years after the SPECT scan, whereas the patient with CAC in the non-proximal LCX remained free of MACE (3-year follow-up). The bottom panel shows two female patients. The patient with CAC in the proximal LCX was younger and had a lower CAC score. She had MACE 5.1 years after the PET scan. The older patient with CAC in the non-proximal LAD remained free of MACE (3 years).

**Figure 2.**
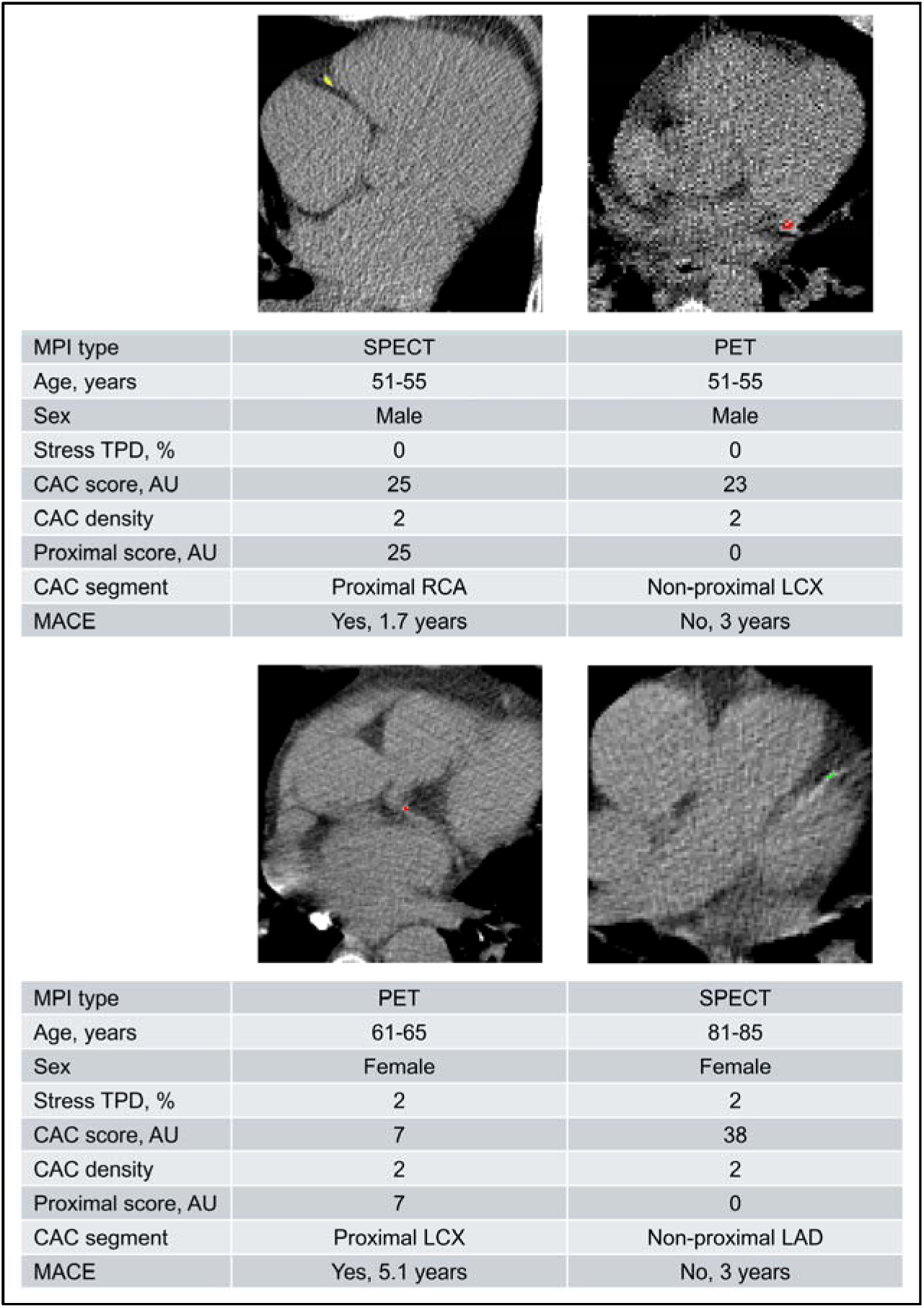
Illustration of proximal involvement and outcome Two pairs of patient cases are presented. The lesion in the left anterior descending artery (LAD) is shown in green, the left circumflex artery (LCX) lesions in red, and the right coronary artery (RCA) lesion in yellow. AU, Agatston Unit; CAC, coronary artery calcium; MACE, major adverse cardiovascular event; MPI, myocardial perfusion imaging; TPD, total perfusion deficit.

## Discussion

In this international multi-center study, we identified CAC in proximal coronary segments from low-dose non-contrast ungated CTAC scans using an AI approach and then evaluated its prognostic importance. We demonstrated the feasibility of a fully automated pipeline for segmenting total CAC and identifying proximal CAC on CTAC scans. We found 9,961 (56%) patients with CAC in proximal coronary segments among 17,830 patients with CAC. In patients with mild CAC scores and normal perfusion, proximal CAC was independently associated with an increased risk of MACE and ACM. Proximal CAC improved risk stratification beyond CAC score and density and other clinical risk factors, providing risk reclassification, particularly for patients without proximal CAC. Our results suggest that AI-enabled identification of proximal CAC could provide additional prognostic value for this patient group.

This is the first study to identify CAC in proximal coronary segments from low-dose non-contrast ungated CTAC scans and to evaluate the prognostic utility of proximal calcification in patients with suspected CAD, specifically those with mild CAC and normal perfusion. In this group from our international cohort, 37% of the patients had proximal CAC identified by AI. These patients were significantly older, more likely to have dyslipidemia, but were less likely to have higher BMI and diabetes. As in previous studies [23, 30], proximal CAC stratified the risk for MACE in the mild-CAC population. We also showed that proximal CAC had a similar stratification power for ACM risk. Patients with proximal CAC had higher cumulative event rates of MACE and ACM than those without proximal CAC. In addition, we found that in this group, patients with proximal CAC had significantly higher total CAC scores and density. After adjusting for CAC score and density[44, 49], the multivariable Cox regression models showed that proximal involvement was associated with a 1.3-fold increase in MACE risk and a 1.4-fold increase in ACM risk. The hazard ratios remained significant after additional adjustment for demographic information, clinical risk factors, and stress perfusion deficit. These suggest that proximal CAC is an independent and complementary predictor for MACE and ACM in this population.

An important implication from our study was that the absence of proximal CAC may help identify a lower-risk subgroup among patients with normal perfusion and mild CAC. This patient population represents a unique challenge for clinical management. Abnormal perfusion is frequently used to guide revascularization decisions and may help target medical therapies for CAD [50]. With respect to initiating medical therapy, statin therapy is generally considered indicated in patients with CAC > 100 [51]. However, among patients with CAC 1-100, the need for this therapy is less clear, with a number needed to treat of 100, compared with 12 among patients with CAC >100 [52]. In addition, CAC 1-99 showed no prognostic difference for ACM compared to CAC=0 in patients who underwent PET scans, particularly in the normal perfusion group. [4] Assuming that the risk of MACE and ACM is proportional to the benefit of statin therapy, the presence or absence of proximal CAC in patients with mild CAC scores may help identify patients more likely to benefit from preventive cardiovascular therapies.

We applied our fully automated pipeline to quantify proximal CAC from CTAC scans without comparing its accuracy against expert annotations. AI identification and measurement may differ from clinicians’ readings. However, the pipeline achieved high agreement with experts in the training dataset during the algorithm development. Additionally, classification of proximal CAC from CTAC is not routinely applied, and therefore, there is no clear ground truth. The use of multi-center scans with multiple imaging protocols makes results more robust and generalizable but may limit precision in estimating associated risk. We only studied the prognostic value of proximal CAC in patients with mild CAC, as this group had been shown to have additional prognostic information for asymptomatic adults. Lastly, the retrospective design of the two registries limits our findings to correlation rather than causation, and future prospective studies are warranted.

## Conclusion

Proximal CAC on CT attenuation maps can be identified using an AI pipeline and is associated with MACE and ACM in patients with suspected CAD, specifically those with mild CAC and normal perfusion. The identification may improve risk stratification for these patients.

## List of Abbreviations

ACM: all-cause mortality
AI: artificial intelligence
CAC: coronary artery calcium
CTAC: computed tomography for attenuation correction
MACE: major adverse cardiovascular event
MPI: myocardial perfusion imaging
PET: positron emission tomography
SPECT: single-photon emission computed tomography

## Statements and Declarations

### Author Contributions

J.Z. and P.J.S. designed the study. G.R., V.B., M.L., R.J.H.M., P.B.K., J.X.L., A.F., E.J.M., A.J.S., T.D.R., L.S., M.I.T., E.A., I.C-J., R.R.S.P., M.A.-M., A.J.E., W.A., S.K., V.T.L., S.M., T.L.R., S.W., P.C., R.A.d., R.R.B., D.S.B., M.F.D.C. and P.J.S. collected and organized data. J.Z., A.S., G.R., V.B., M.L., A.K., R.J.H.M., P.B.K. and J.X.L. curated the dataset. J.Z. developed the AI prediction code, performed statistical tests, and created all figures. J.Z., R.J.H.M. and P.J.S. drafted the manuscript with feedback from all authors. P.J.S. supervised the study. All authors discussed the results and approved the final version before submission.

### Data Availability

To the extent allowed by data sharing agreements and IRB protocols, data from this manuscript will be shared upon written request.

### Ethics Approval

The overall registries were approved by the investigational review board at Cedars-Sinai Medical Center and complied with the Declaration of Helsinki.

### Consent to participate

Informed consent was obtained from all individual participants included in the study.

### Consent to publish

The authors affirm that human research participants provided informed consent for publication of the images in Figure 2 and Graphical Abstract.

