## Supplemental Material for "Hidden risk in normal myocardial perfusion scans: AI-detected proximal coronary calcium on CT attenuation maps improves prognosis"

Supplement Figure 1. Study population

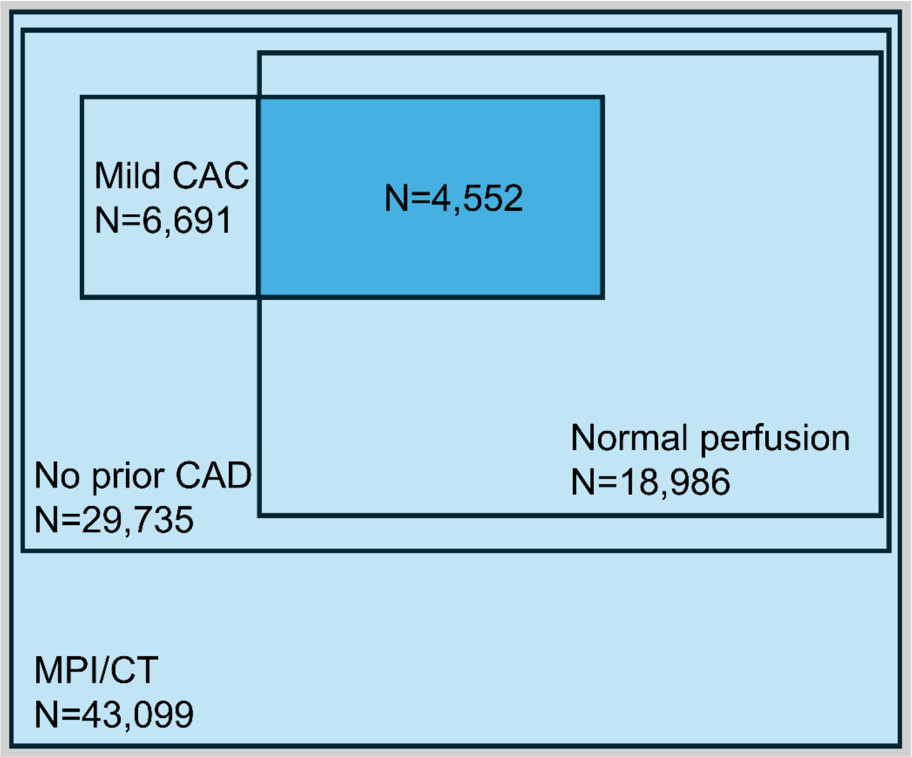


From hybrid myocardial perfusion imaging (MPI)/CT (N=43,099), the study sample (N=4,552) was selected for 1) no prior coronary artery disease (CAD), 2) mild coronary artery calcium (CAC), and 3) normal perfusion.

Supplement Table 1. CT acquisition protocols with SPECT imaging

| Site | Number of patients | Scanner system | Slice thickness, mm | Tube current, mA | Tube voltage, kVp |
|---|---|---|---|---|---|
| University of Calgary | 2,912 | GE Discovery 570 | 5 | 20 | 120 |
| Columbia University | 1,878 | Philips Precedence 16P | 3 | 30 | 120 |
| Ottawa Heart Institute | 1,433 | Siemens Symbia Intevo | 5 | 20-30 | 130 |
| Yale University | 4,766 | GE Discovery 570 | 2.5 | 60 or 150, based on body-mass index | 120 |

Supplement Table 2. CT acquisition protocols with PET imaging

| Site | Number of patients | Scanner system | Slice thickness, mm | Tube current, mA | Tube voltage, kVp |
|---|---|---|---|---|---|
| Brigham and Women's Hospital | 7,200 | GE Discovery MI<br>GE Discovery MI RX<br>GE Discovery MI STE | 2.5-5 | 10-26 | 120-140 |
| Cedars-Sinai Medical Center | 5,002 | Siemens Biograph 64 TruePoint<br>Siemens Biograph 128 Vision Edge<br>GE Discovery 710 | 3 | 11-13 | 100 |
| Columbia University Irving Medical Center | 1,922 | Siemens Biograph 64 mCT Flow | 3 | 75 | 120 |
| Houston Methodist Academic Institute | 1,876 | Siemens Biograph 600 Vision Edge | 3 | 20-50 | 100-120 |
| Intermountain Medical Center | 5,907 | Siemens Biograph 16 TruePoint<br>Siemens Biograph 20 mCT<br>Siemens Biograph 40 mCT | 2 | 15-38 | 120-130 |
| University of Kansas Medical Center | 589 | GE Discovery MI | 3.75 | 14-75 | 120 |
| Montefiore Medical Center | 1,014 | Philips Gemini TF TOF 16 | 3 | 110-185 | 120 |

| | | | | | |
|---|---|---|---|---|---|
| | | Philips Gemini TF TOF 64 | | | |
| Mayo Clinic | 1,989 | GE Discovery 710 | 3.75 | 17-77 | 120 |
| National Autonomous University of Mexico | 323 | Siemens Biograph 64 TruePoint<br>Siemens Biograph Vision 600 | 3 | 33-580 | 120 |
| University of Naples Federico II | 455 | Philips Ingenuity TF<br>GE Discovery Mi | 3 | 35-100 | 120-140 |
| Ottawa Heart Institute | 3,934 | GE Discovery 690<br>GE Discovery 600<br>Siemens Biograph 600 Vision Edge | 3-5 | 20-60 | 120 |
| West Los Angeles Veterans Affairs Medical Center | 1,150 | Siemens Biograph 64 mCT<br>Siemens Biograph 64 Vision 600 | 3 | 70-200 | 120 |
| University Hospital Zurich | 749 | GE Discovery STE<br>GE Discovery RX<br>GE Discovery LS<br>GE Discovery HR | 3.75-5 | 140 | 40-240 |

Supplement Table 3. Likelihood ratio test and continuous net reclassification analysis.

| Outcome | Base C-index | Nested C-index | Likelihood ratio test P-value | NRI (95% CI) |
|---|---|---|---|---|
| MACE | 0.645 ± 0.013 | 0.648 ± 0.013 | 0.020 | 12% (2%-19%) |
| ACM | 0.680 ± 0.015 | 0.683 ± 0.014 | 0.037 | 12% (1%-24%) |

C-indices were shown with standard deviations. Base model: adjustment 4 in Table 2; nested model: base model + proximal involvement. ACM, all-cause mortality; CI, confidence interval; MACE, major adverse cardiovascular events; NRI, net reclassification index.
